# Spinal Cord Motor Neuron Phenotypes and Polygenic Risk Scores in Sporadic Amyotrophic Lateral Sclerosis: Deciphering the Disease Pathology and Therapeutic Potential of Ropinirole Hydrochloride

**DOI:** 10.1101/2024.03.04.24303652

**Authors:** Chris Kato, Satoru Morimoto, Shinichi Takahashi, Shinichi Namba, Qingbo S. Wang, Yukinori Okada, Hideyuki Okano

**Author notes:** **Co-corresponding author Correspondences to:** Hideyuki Okano, MD, PhD, Dean, Keio University Graduate School of Medicine, Professor, Department of Physiology, Full address: Department of Physiology, Keio University School of Medicine, 35, Shinanomachi, Shinjuku-ku, Tokyo, 160-8582, Japan, Tell, Satoru Morimoto, MD, PhD, Instructor, Department of Physiology, Department of Physiology, Keio University School of Medicine, 35 Shinanomachi, Shinjuku-ku, Tokyo, 160-8582, Japan, Tell.

## Abstract

This study investigated the genetic factors associated with the onset of amyotrophic lateral sclerosis (ALS) and the mechanism of action of Ropinirole hydrochloride (ROPI) in ALS treatment. Firstly, ALS is a neurodegenerative disease characterized by the degeneration of both upper and lower motor neurons (MNs), and identifying the genetic factors of sporadic ALS (SALS) has been challenging. However, ALS-like phenotypes have been observed in induced pluripotent stem cell-derived lower MNs (iPSC-LMNs) of SALS patients, suggesting a polygenic contribution to SALS pathogenesis, and ROPI has emerged as a novel therapeutic candidate for ALS treatment. Moreover, recent genome-wide association studies (GWAS) have suggested an association between high levels of blood total cholesterol (bTC) and ALS. The study aimed to combine iPSC-LMN phenotypes and polygenic risk scores (PRS) to quantify the genetic background of patients and analyze the mechanism of action of ROPI in ALS treatment. Twenty SALS patients were included in the study, and iPSC-LMN phenotypes, PRS for 58 quantitative traits, and ALS were calculated. The results indicated that patients with a higher PRS for bTC showed changes in iPSC-LMN phenotypes, particularly a higher response to ROPI. Additionally, a correlation was observed between bTC levels and the expression of cholesterol biosynthesis (CB) enzymes in MNs, suggesting their involvement in the pathogenesis of SALS. Furthermore, the study compared the liver and spinal cord expression of CB enzymes and found a correlation between bTC levels and CB enzyme expression, particularly in MNs, which may partly explain the MN-specific pathogenesis of SALS. In conclusion, the study suggests that high bTC levels and increased CB activity may contribute to the pathogenesis of SALS, and CB suppression may be a potential therapeutic strategy for ALS, supported by the mechanism of action of ROPI in ALS treatment.

## INTRODUCTION

Amyotrophic lateral sclerosis (ALS) is a neurodegenerative disease characterized by both upper and lower motor neuron (MN) degeneration. Approximately 90% of ALS patients have sporadic ALS (SALS). Thus, identifying the genetic factors of SALS is difficult. However, lower MNs (LMNs), including spinal MNs, that derive from induced pluripotent stem cells (iPSC-LMNs) in SALS patients exhibit ALS-like phenotypes,^1^ such as reduced neurite length (NL), suggesting a polygenic contribution to SALS pathogenesis.

Drug screening using iPSC-LMNs recently identified ropinirole hydrochloride (ROPI) as a novel therapeutic candidate for ALS.^1^ A clinical trial of ROPI (the Ropinirole Hydrochloride Remedy for Amyotrophic Lateral Sclerosis [ROPALS] trial) involving 20 SALS patients suggested that ROPI inhibits the pathological progression in ALS development.^2^ Furthermore, our reverse translational study in the ROPALS trial revealed that patients who had iPSC-LMNs that responded strongly to ROPI showed greater clinical efficacy in the trial.^2^ This suggests that iPSC-MN phenotypes reflect drug responsiveness in SALS patients.

Recent genome-wide association studies (GWASs) have uncovered genetic predispositions for various traits in individuals. A recent GWAS in ALS patients suggested that the mutations associated with high levels of blood total cholesterol (bTC) play a causal role in ALS development.^3^ The polygenic risk score (PRS) is a quantitative measure that is obtained by summing the small effects of each risk variant identified by GWASs and is widely used to predict diverse traits in individuals.

In the present study, we aimed to identify the genetic factors in the development of SALS and the mechanism underlying the anti-ALS effect of ROPI by combining two indices that reflect genetic predisposition: the phenotype of iPSC-LMNs and the PRS (Fig. 1A).

**Figure 1.**
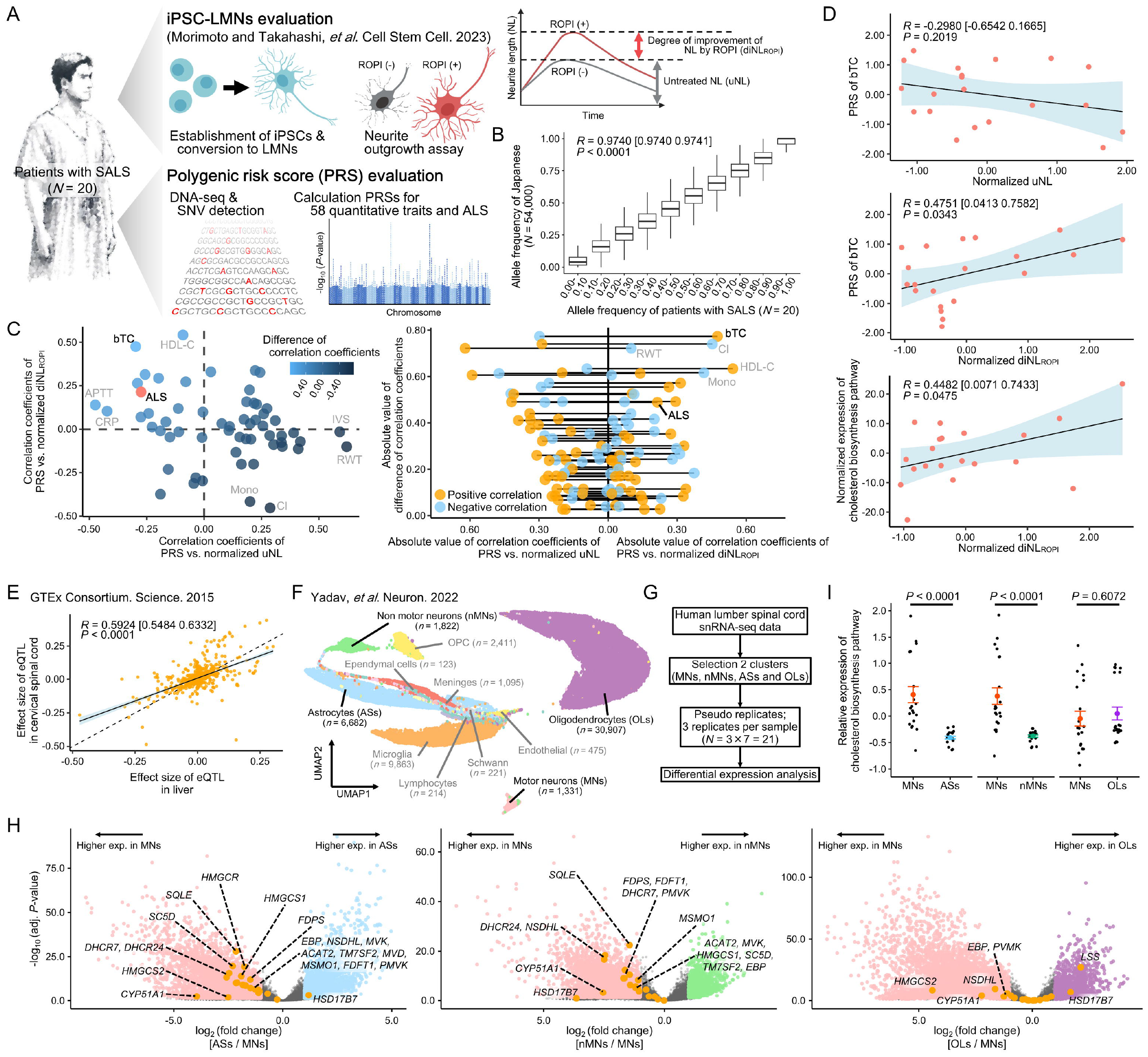
Analysis of lower motor neuron derived from induced pluripotent stem cell (iPSC-LMN) phenotypes and polygenic risk scores (PRSs) in sporadic amyotrophic lateral sclerosis (SALS) patients revealed that increased cholesterol biosynthesis (CB) in LMNs is vital to SALS pathology, and the suppression of CB by ropinirole hydrochloride (ROPI) is a key treatment strategy. (**A**) Schematic diagram of the study. (**B**) Comparison of allele frequencies between SALS patients from the Ropinirole hydrochloride remedy for amyotrophic lateral sclerosis (ROPALS) trial (*N* = 20) and the Japanese allele frequency panel (Tohoku Medical Megabank Organization [ToMMo], 54KJPN). (**C**) **(Left)** Results of the Pearson’s correlation analyses between the PRSs for 58 quantitative traits and the iPSC-LMN phenotypes (untreated neurite length [uNL] and degree of improvement in neurite length by ROPI [diNL_ROPI_]). Each plot represents the correlation coefficient (amyotrophic lateral sclerosis [ALS]: salmon pink, 58 quantitative traits: blue). Differences in correlation coefficients between the PRSs (for ALS and 58 quantitative traits) and iPSC-LMN phenotypes (uNL and diNL_ROPI_; which reflects the extent of ALS pathology and the degree of improvement following ROPI administration, respectively) are shown in shades of blue (the brighter the trait, the more ALS pathology is exacerbated by high PRS for that trait while it improves with ROPI administration). **(Right)** Differences in correlation coefficients between the PRSs (for ALS and 58 quantitative traits) and iPSC-LMN phenotypes (uNL and diNL_ROPI_) are shown on the vertical axis, and the absolute values of each correlation coefficient are shown on the horizontal axis. The left half shows the absolute values of the correlation coefficients for uNL, and the right half shows those for diNL_ROPI_. Orange circles indicate traits with positive correlations, and light blue circles indicate traits with negative correlations. (**D**) Results of the Pearson’s correlation analyses between the PRS for blood total cholesterol (bTC) and iPSC-LMN phenotypes (uNL and diNL_ROPI_; upper and middle graph). Results of the Pearson’s correlation analysis between CB enzyme expression in iPSC-MNs and its diNL_ROPI_ (lower graph). (**E**) Comparison of tissue-specific effect sizes of the liver and cervical spinal cord (SC) for expression quantitative trait loci (eQTLs) visualizing the expression levels of CB enzymes (Pearson’s correlation test). Each plot represents each eQTL. (**F**) Uniform manifold approximation and projection (UMAP) of human lumber SC cells. We used the same cell-type identification method as that used in the original paper. (**G**) Overview of pseudo-bulk differential expression analysis. (**H**) Volcano plots showing the results of the pseudo-bulk differential expression analysis, compared by cell type (MNs, astrocytes [ASs], non-MNs [nMNs], and oligodendrocytes [OLs]). From left to right, results for ASs vs. MNs, nMNs vs. MNs, and OLs vs. MNs are presented. Fold changes are calculated relative to MNs. (**I**) Graphs showing relative expression of CB pathway enzymes by cell-type comparisons (MNs, ASs, nMNs, and OLs). Each plot represents the expression index of CB enzymes per pseudo-bulk. Coloured plots represent the mean expression index for each cell type. Error bars represent standard errors of the mean. SALS, sporadic amyotrophic lateral sclerosis; iPSC-LMN, lower motor neuron derived from induced pluripotent stem cell; ROPI, ropinirole hydrochloride; NL, neurite length; diNL_ROPI_, degree of improvement of NL by ROPI; uNL, untreated NL; PRS, polygenic risk score; DNA-seq, deoxyribose nucleic acid sequence; SNV, single nucleotide variant; adj. *P*-value, adjusted *P*-value; bTC, blood total cholesterol; HDL-C, high-density-lipoprotein cholesterol; APTT, activated partial thromboplastin time; CRP, C-reactive protein; ALS, amyotrophic lateral sclerosis; Mono, monocyte count; Cl, chloride; IVS, interventricular septum thickness (echocardiographic); RWT, relative wall thickness (echocardiographic); vs., versus; GTEx, genotype-tissue expression; eQTL, expression quantitative trait loci; OPC, oligodendrocyte precursor cell; UMAP, uniform manifold approximation and projection; RNA, ribonucleic acid; snRNA-seq, single-nucleus RNA sequence; ASs, astrocytes; OLs, oligodendrocytes; exp., expression.

## METHODS

This study included 20 SALS patients who participated in the ROPALS trial. We used a dataset obtained for the ROPALS trial,^2^ which involved measuring the NL and the transcriptome of iPSC-LMNs with and without ROPI administration (Fig. 1A). We calculated both the untreated NL (uNL) and the degree of improvement of NL by ROPI (diNL_ROPI_). Additionally, we calculated PRSs for 58 quantitative traits and ALS (Supplementary Table 1). Using public data, we also analyzed the liver-cervical spinal cord (SC) comparison of expression quantitative trait loci (eQTLs) effect sizes, and the transcriptome in various cell types in the lumbar SC.

The public data sources are detailed in Supplementary Table 2. More detailed methodological information is provided in the Supplementary Methods.

## RESULTS

The allele frequencies of SALS patients in the ROPALS trial were highly similar to the Japanese allele frequency panel (*P* < 0.0001, *R* = 0.9740 [0.9740 0.9741]), which confirmed appropriate sequence determination (Fig. 1B). Moreover, the iPSC-LMN phenotypes (i.e., uNL and diNL_ROPI_) showed a weak non-significant correlation with the PRS of ALS, which suggests that the phenotypes reflect the genetic background in ALS (Supplementary Fig. 1A).

The correlation analysis between iPSC-LMN phenotypes and the PRSs for 58 quantitative traits and ALS revealed that the greatest difference in correlation coefficients between uNL and diNL_ROPI_ was observed in bTC (Figure 1C; Supplementary Table 1). Patients with a higher bTC PRS tended to have shorter uNL (*P* = 0.2019, *R* = −0.2980 [−0.6542 0.1665]) and greater diNL_ROPI_ (*P* = 0.0343, *R* = 0.4751 [0.0413 0.7582]; Fig. 1D). Moreover, patients with higher expression of cholesterol biosynthesis (CB) enzymes (Supplementary Table 3) in iPSC-LMNs had greater diNL_ROPI_ (*P* = 0.0475, *R* = 0.4482 [0.0071 0.7433]; Fig. 1D).

bTC levels generally reflect CB levels in the liver, whereas cholesterol levels in the SC are independent of systemic circulation. Therefore, we compared the sizes of the effect of eQTLs on CB enzyme expression in the liver and cervical SC using the genotype-tissue expression project data, which showed high conservation (*P* < 0.0001, *R* = 0.5924 [0.5484 0.6332]; Fig. 1E).

The cell-type specificity of CB in the SC was examined using pseudo-bulk differential expression analysis, which was based on public single-nucleus ribonucleic acid sequencing data of the human lumbar SC (Fig. 1F and G). The marker genes of each cell type confirmed that the cell types were appropriately identified (Supplementary Fig. 1B). Results showed that CB enzyme expression in MNs was higher than that in astrocytes (ASs) or non-MNs (nMNs) and similar to that in oligodendrocytes (OLs; MNs vs. ASs: *P* < 0.0001, vs. nMNs: *P* < 0.0001, vs. OLs: *P* = 0.6072; Fig. 1H and I).

## DISCUSSION

Our findings suggested that individuals with a genetic predisposition for a high level of bTC may also have increased CB activity in the liver, SC, and ultimately MNs, which may result in an ALS-like phenotype of reduced NL in iPSC-LMNs.

Given that the genetic factors associated with high bTC levels play a causal role in the development of SALS, as reported by a large-scale GWAS in ALS patients,^3^ it is plausible that CB hyperactivity in MNs (determined by genetic background) contributes to the development of SALS. It has previously been reported that CB hyperactivity increases the presence of cholesterol esters, a neurotoxic metabolite, and this has been proposed as a mechanism underlying MN death in ALS.^4^ Indeed, elevated levels of cholesterol and cholesterol esters have been observed in the spinal fluid of SALS patients.^4,5^ Furthermore, increased CB activity in the mouse SC has been shown to produce ALS-like symptoms.^4^ The enhanced CB activity in MNs compared with nMNs suggests cell-type specificity, which could partly explain the MN-specific pathogenesis of SALS.

In contrast, a higher PRS for bTC was associated with a better response to ROPI *in vitro*. Because ROPI has been shown to suppress CB enzyme expression in iPSC-LMNs,^2^ the suppression of CB at the MN level may be a potential anti-ALS mechanism of ROPI.

In conclusion, the combined analysis of iPSC-LMN phenotypes and PRSs to quantify the genetic background of patients showed that increased CB in MNs is pivotal for SALS pathology and its suppression may be the mechanism underlying the anti-ALS effect of ROPI. Thus, CB suppression may be a viable strategy for the treatment of SALS.

## Data Availability

All data produced in the present study are available upon reasonable request to the authors.

## Acknowledgements

We thank the coordinators, staff, physicians, and participants of the ROPALS trial. We are also grateful to Shiho Nakamura, Fumiko Ozawa, Naoki Kobayashi, Hiroya Kobayashi, Yuki Oguma (Keio University), and Selena Setsu (University of Tokyo). We extend our special thanks to Professor Hiroshi Nishihara (Genomics Unit, Keio Cancer Center, Keio University Hospital) for his invaluable advice on genomic analysis.

## Contributors

C.K., S.M. and S.T. designed the study and acquired samples from patients. C.K. analysed the data and performed statistical analyses. C.K., S.M., S.T., S.N., Q.S.W., Y.O. and H.O. contributed to the interpretation of the results. S.T., S.N., Q.S.W., Y.O. and H.O. provided supervision. C.K. and S.M. wrote the manuscript, and all authors corrected the manuscript and collectively made the decision to submit for publication.

## Patient consent for publication

Not applicable.

## Ethics approval

This study was approved by the Institutional Review Board of Keio University School of Medicine (approval No. 20080016), and written informed consent was obtained from the participants.

