## Supplementary for "Spinal Cord Motor Neuron Phenotypes and Polygenic Risk Scores in Sporadic Amyotrophic Lateral Sclerosis: Deciphering the Disease Pathology and Therapeutic Potential of Ropinirole Hydrochloride": Kato_et_al_medRxiv_supple document.docx

**SUPPLEMENTARY MATERIALS**

**Supplementary Methods**

***Participants***

We included 20 SALS patients who participated in the ROPALS trial. Detailed patient information and inclusion/exclusion criteria are provided in the trial report.^1^

***SNP detection and PRS calculation***

Whole genome sequencing was performed in 20 patients with SALS who participated in the ROPALS trial (Fig. 1A). For quality control, all fastq files were pre-processed using Trimmomatic (version 0.39; ILLUMINACLIP:TruSeq3-PE-. 2.fa:2:30:10 LEADING:30 TRAILING:30 SLIDINGWINDOW:5:30 MINLEN:75). Following quality control, the fastq files were mapped to the hg19 reference sequence by bwa mem in BWA (version 0.7.17-r1188),^2^ converted into binary alignment and map (BAM) files using samtools (version 1.17)^3^ and marked for duplicates using MarkDuplicates in picard (version 3.0.0).^4^ Subsequently, base quality recalibration was performed using BaseRecalibrator in gatk (version 4.0.0.0).^5^ Finally, variants were identified using HaplotypeCaller in gatk and variant call format (VCF) files were obtained by GenotypeGVCFs in gatk. Single nucleotide polymorphism (SNP)-only VCF files were also obtained using SelectVariants in gatk, and filtered using VariantFiltration in gatk (-filter 'QD < 2.0' --filter-name 'QD2' -filter 'QUAL < 30.0' --filter- name 'QUAL30' -filter 'SOR > 4.0' --filter-name 'SOR4' -filter 'FS > 60.0' --filter-name 'FS60' -filter 'MQ < 40.0' --filter-name 'MQ40'). For the calculation of the PRS, summary statistic of ALS was obtained from Rheenen W, et al.^6^ and of 58 quantitative traits were obtained from Kanai M, *et al*.^7^, and PRScs (version 1.1.0, the UK biobank EAS reference as LD reference)^8^ was used for the calculation.

***Pseudo-bulk differential expression analysis***

The R package Seurat (version 4.3.0)^9^ was used for the analysis of single-nucleus ribonucleic acid sequencing (snRNA-seq) data, and umap (version 0.2.10.)^10^ was used for the uniform manifold approximation and projection (UMAP) analysis. We obtained the count data of the snRNA-seq analysis, and the annotation of cell types was based on previous study.^11^ Because the data used were samples acquired from seven individuals, we randomly split the data so that *n* = 3 per individual and per cell type (seed = 123) to obtain the pseudo-bulk data. DESeq2 (version 1.34.0)^12^ was used for the comparison between the two groups, with a cut-off *P*-value of 0.05 and a cut-off log_2_(fold change) of ±1.

The relative expression of the CB pathway was determined by performing z-normalization for each gene listed in Supplementary Table 1 on the post-split pseudo-bulk data, followed by the summation of the relative expression levels of all genes.

***Statistical analysis and other analysis tools***

All statistical analyses were performed using R (version 4.1.2).^13^ To calculate the correlation coefficients and 95% confidence intervals, we used the R package psych (version 2.3.6).^14^ Student's t-tests were used to compare continuous data between the two groups. The cut-off value for the *P*-value was 0.05, and all tests were two-tailed. All visualizations were performed using the R package, ggplot2 (version 3.4.3).^15^

**Supplementary Results and Discussion**

Correlation analysis was performed between the PRS for ALS, and uNL and diNL_ROPI_ as phenotypes of iPSC-LMNs (Supplementary Fig. 1A). Results showed a weak non-significant negative correlation for uNL and a weak non-significant positive correlation for diNL_ROPI_ (Supplementary Fig. 1A, uNL: *P* = 0.2413, *R* = −0.2746 [−0.6394 0.1911], diNL_ROPI_: *P* = 0.3648, *R* = 0.2140 [−0.2524 0.5998]). These results are important as proof of concept because they suggest that iPSC-LMNs partially reflect the genetic background of ALS. However, the correlation was weaker than that observed in bTC and some other traits (Figs. 1C, 1D).

There are several hypotheses as for why the PRSs for bTC and some other traits reflect the phenotypes uNL and diNL_ROPI_ more than the PRS for ALS. The first is that the pathogenesis of ALS can also be observed in cells other than LMNs, including upper MNs. Because the only cells we evaluated as models of ALS pathogenesis were LMNs only, they may not reflect the full genetic background of ALS. Thus, it will be important to reproduce conditions closer to the disease environment by generating various cell types from iPSCs, such as organoids, which have been studied recently.^16^ Nevertheless, our results showing that the PRS for bTC mirrored the phenotypes uNL and diNL_ROPI_ more closely than the PRS for ALS suggest that the pathology in LMNs can be partly explained by the over activation of CB. Second, although the PRS for bTC (and other traits) was calculated for the quantitative trait (i.e., bTC level), the PRS for ALS was calculated for the discrete trait of disease presence (i.e., ALS or healthy). Patients with ALS are highly variable in terms of variety of factors, including age of onset and speed of symptom progression. Therefore, it is unclear what clinical traits (e.g., age of onset) the size of the PRS for ALS reflects. These hypotheses need to be elucidated through more detailed analyses in the future.

We compared the allele frequencies of the mutations involved in the pathogenesis of ALS between the 20 patients with SALS who participated in the ROPALS trial and the Japanese allele frequency panel. Candidate mutations involved in the pathogenesis of ALS identified by summary statistics of GWAS in ALS patients were narrowed down using a cut-off of *P* < 5×10^−8^, and the difference in frequency between the SALS patients in the ROPALS trial and the Japanese allele frequency panel, and the effect sizes of these mutations were then plotted (Supplementary Fig. 1C). Results showed that mutations with a risk of developing ALS were more common in the Japanese panel, whereas mutations that had a protective effect on ALS development were more common in the SALS patients in ROPALS trial. This contradictory result may be attributed to our small sample size. Mutations with low frequencies in the Japanese population are unlikely to be found in the SALS patients in the ROPALS trial, which would result in an apparently lower frequency of rare mutations in the SALS patients. Mutations at risk of developing ALS are uncommon; thus, there is a low frequency of mutations at risk of developing ALS in the SALS patients in the ROPALS trial (Supplementary Fig. 1C). Therefore, we considered that the calculation of the PRS for ALS in each patient would provide a more comprehensive picture of the genetic background of ALS development and that our PRS calculation was valid.

Previously, the main CB producing cells in the central nervous system (CNS) were considered to be OLs and ASs, and CB is thought to be suppressed in mature neurons, which take up cholesterol from glial cells such as ASs.^17–19^ To our knowledge, to date, cell-type heterogeneity of cholesterol metabolism in neurons, including MNs, has not been investigated. In our study, we used public snRNA-seq data to divide neurons into two groups: MNs and nMNs. Intriguingly, the results showed that CB was more active in MNs than in ASs and nMNs but occurred similarly to that in OLs. OLs are a cell type that synthesizes large amounts of cholesterol to form the myelin sheath.^19^ Thus, it is of interest that the level of CB in MNs was comparable to that in OLs. The physiological activation of CB in MNs may result in the production of large amounts of potentially neurotoxic cholesterol metabolites.^20–22^ Therefore, it has been hypothesized that the higher risk of developing ALS in individuals with a genetic background of high bTC is due to the pathological activation of CB in MNs and a breakdown of homeostasis owing to an inability to process cholesterol metabolites and other factors.

On the other hand, ROPI, known for suppressing the expression of CB enzymes,^23^ demonstrated enhanced effectiveness *in vitro*, particularly in patients with a genetic predisposition to high bTC and elevated CB enzyme expression in iPSC-LMNs. These results suggest that the mechanism of action of ROPI may involve the inhibition of CB in iPSC-LMNs, as discussed in the Discussion section.

In our earlier study on drug screen using ALS patient-derived iPSC-LMNs as a model for ALS pathology, we identified nine candidate drugs.^24^ Notably, one of these candidates, alendronate sodium, acts as an inhibitor of farnesyl pyrophosphate synthase (FPPS) within the mevalonate pathway, a part of the CB pathway.^25–27^ Thus, ROPI and alendronate sodium appear to share a common function in inhibiting CB, as the shared mechanism of the anti-ALS effect. Moreover, cortisone acetate, ranked as the second most effective candidate after ROPI, functions as a glucocorticoid receptor agonist and is commonly administrated as an anti-inflammatory agent. Among the 58 quantitative traits, the second trait correlating with ALS-like phenotype in iPSC-LMNs (i.e., uNL) was the blood level of C-reactive protein (CRP), a blood marker elevated during inflammation. In general, the anti-inflammatory drug glucocorticoids, including cortisone acetate reduce blood CRP levels (one should be cautious about generalizing its effects inside and outside the CNS, because CRP is synthesized primary in the liver),^28,29^ highlighting the possibility that the traits identified by exploratory correlation analysis of the iPSC-LMN phenotype and the PRS may be useful as therapeutic targets. Therefore, while ROPI may targets CB in the LMNs for its anti-ALS effect, drugs influencing other traits, such as active partial thromboplastin time (APTT) and CRP, may also serve as viable therapeutic targets. A more comprehensive ALS therapeutic strategy might be achieved by targeting multiple therapeutic goals with various agents, including ROPI and alendronate acetate. Comparative analyses of the iPSC-LMN phenotype and the PRS could thus play a pivotal role not only in elucidating disease mechanisms but also in identifying new therapeutic targets and exploring potential combination therapies.

**Supplementary limitations**

One limitation of our study is the relatively small sample size of 20 participants. Thus, larger-scale studies are warranted. Additionally, we did not biochemically measure the CB levels in iPSC-LMNs. Furthermore, it remains unclear how the upregulation of CB contributes to the shortening of neurite length in iPSC-MNs and the onset of ALS. This highlights the need for further research in this area.

**Supplementary figure legend**

**(A)** Results of Pearson’s correlation analyses of the PRS for ALS and iPSC-LMNs phenotypes (uNL and diNL_ROPI_; upper and middle).

**(B)** Volcano plots showing the results of pseudo-bulk differential expression analysis, compared by cell type (MNs, ASs, nMNs, and OLs). Representative genetic markers in MNs, OLs and ASs are shown in deep colours. From upper left, upper right and lower left, results for ASs vs MNs, nMNs vs MNs and OLs vs MNs are presented. Fold changes are calculated relative to MNs.

**(C)** Scatter plot showing the comparison of allele frequencies of candidate mutations between Japanese allele frequency panel and 20 patients with SALS. Candidate mutations were identified through GWAS in ALS patients and filtered using a significance threshold of P < 5×10^−8^. Each point represents a mutation, with its position reflecting the allele frequency difference between the SALS patients in ROPALS trial and the Japanese panel, and its size indicating the effect size. The frequencies in the Japanese allele frequency panel are indicated by the colour intensity, and the *P*-value for each mutation is indicated by the plot size.

PRS, polygenic risk score; ALS, amyotrophic lateral sclerosis; NL, neurite length; uNL, untreated NL; ROPI, ropinirole hydrochloride; diNL_ROPI_, degree of improvement of NL by ROPI; adj. *P*-value, adjusted *P*-value; MNs, motor neurons; ASs, astrocytes; OLs, oligodendrocytes; exp., expression; SALS, sporadic ALS.

4.	Picard toolkit. *Broad Institute, GitHub repository*. Published online 2019.

10.	McInnes L, Healy J, Melville J. UMAP: Uniform Manifold Approximation and Projection for Dimension Reduction. Published online February 9, 2018.

**Supplementary table 1. A list of 58 quantitative traits and ALS, and their results of correlation analysis with uNL and diNL_ROPI_**

**Supplementary table 2. Data information**

**Supplementary table 3. A list of enzymes of cholesterol synthesis pathway**
