## Supplementary figures and images for "Spinal Cord Motor Neuron Phenotypes and Polygenic Risk Scores in Sporadic Amyotrophic Lateral Sclerosis: Deciphering the Disease Pathology and Therapeutic Potential of Ropinirole Hydrochloride"

### Kato_et_al_medRxiv_supple figure.png

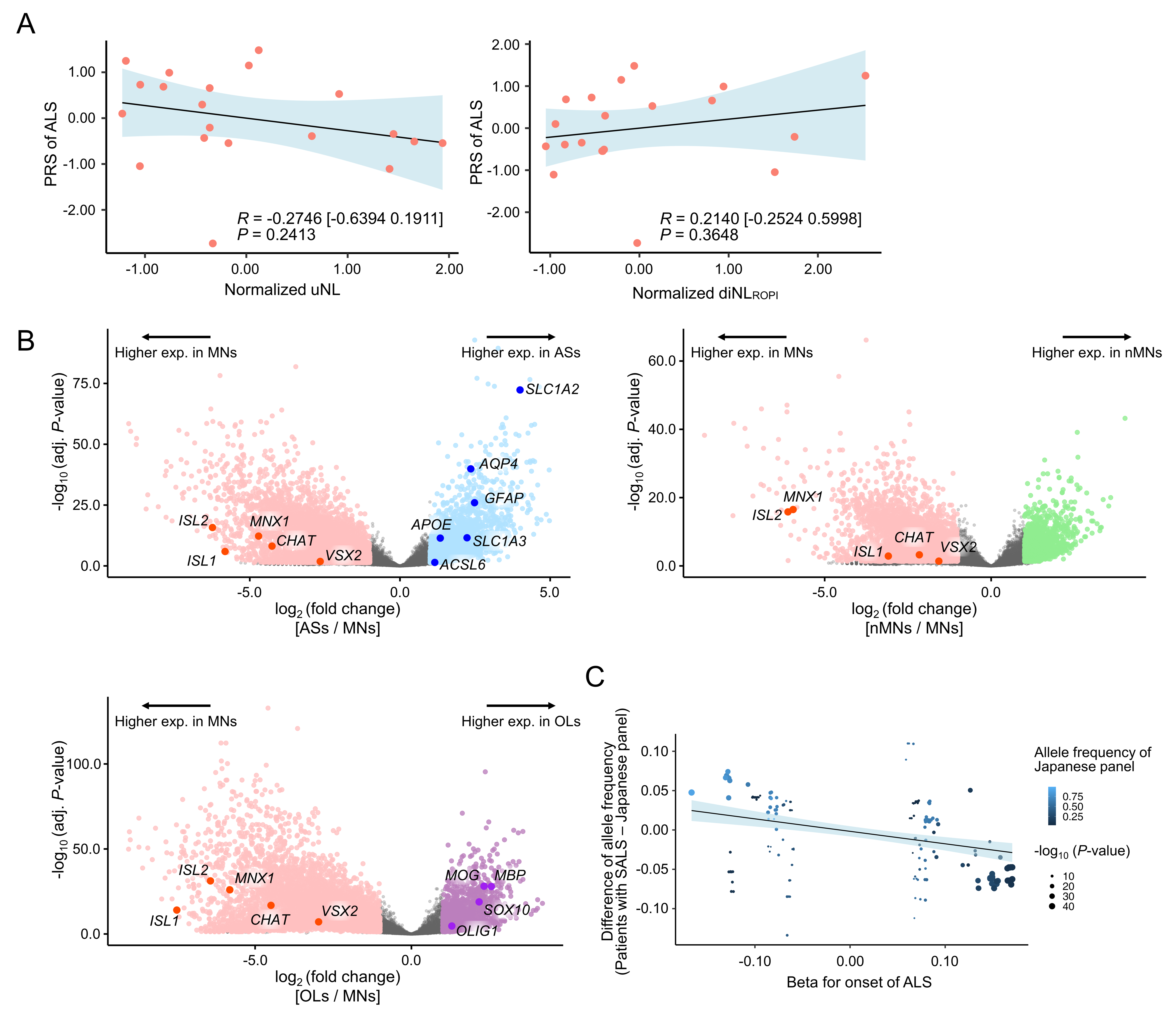
